# Genetic Associations Between Celiac Disease and Type 2 Inflammatory Diseases: A Mendelian Randomization Analysis

**DOI:** 10.1101/2024.01.18.24301488

**Authors:** Mahmud Omar, Mohammad Omar, Saleh Nassar, Adi Lahat, Kassem Sharif

**Affiliations:** Tel-Aviv university faculty of medicine; School of Medicine, V.N.Karazin Kharkiv National University; Edith Wolfson Medical Center; Department of Gastroenterology, Sheba Medical Center, Tel-Hashomer, Israel; Department of Medicine B, Zabludowicz Center for Autoimmune Diseases, Sheba Medical Center, 5262100, Tel-Hashomer, Israel

**Author notes:** **Corresponding author:** Mahmud Omar, Tel-Aviv university, Faculty of medicine. Sheba medical center. **Financial disclosure** – none.

**Keywords:** Celiac Disease, Type 2 Inflammatory Diseases, Mendelian Randomization, Genetic Associations, Asthma, Allergic Rhinitis, Atopic Dermatitis

## Abstract

**Background:** Celiac disease, a gluten-triggered autoimmune disorder, is known for its systemic inflammatory effects. Epidemiological data suggest an association with type 2 inflammatory diseases like asthma, allergic rhinitis, and atopic dermatitis, however, genetic associations remain unclear, prompting this study to explore their potential genetic interplay.

**Methods:** Utilizing Two-Sample Mendelian Randomization (TSMR), we examined genetic associations using 15 genetic instruments from GWAS datasets. Our analysis focused on celiac disease and its relation to asthma, allergic rhinitis, and atopic dermatitis. Power analysis was conducted to determine the study’s detection capabilities, and Odds Ratios (ORs) with 95% confidence intervals (CIs) were calculated using various MR methods.

**Results:** A significant positive association was observed between celiac disease and atopic dermatitis (OR = 1.037, 95% CI: 1.015 - 1.059), and a slight association with allergic rhinitis (OR = 1.002, 95% CI: 1.0004 - 1.0032). Conversely, a minor protective effect was noted for asthma (OR = 0.97, 95% CI: 0.96 - 0.98). These results, supported by a high F-statistic, suggest a strong genetic linkage, despite some heterogeneity and complexity in the associations.

**Conclusion:** Our study identifies significant genetic links between celiac disease and type 2 inflammatory diseases, particularly atopic dermatitis and allergic rhinitis, with a minor protective effect against asthma. These findings, underscored by a strong F-statistic, suggest complex genetic interactions and emphasize the need for further research to explore their clinical relevance.

## Introduction

Celiac disease is an autoimmune disorder triggered by the ingestion of gluten in genetically predisposed individuals, leading to a range of gastrointestinal and systemic symptoms (1,2). It is characterized by inflammation of the small intestine, which can result in malabsorption of nutrients (2). Beyond the gut, celiac disease has been associated with a spectrum of extraintestinal manifestations, suggesting a broader systemic inflammatory response (3). Similarly, type 2 inflammatory diseases, such as asthma, allergic rhinitis, and atopic dermatitis, are characterized by chronic inflammatory processes, often presenting with a dysregulated immune response to environmental factors (4–6).

The connection between celiac disease and type 2 inflammatory diseases has been a subject of interest due to overlapping clinical features and shared inflammatory pathways, such as T-helper cell type 2 (Th2) mediated immune responses (6–8). These pathways involve cytokines and other inflammatory mediators that play a role in both the pathogenesis of celiac disease and allergic diseases (2,8). However, the exact nature of the genetic and molecular interplay between these conditions remains inadequately defined (9).

Two-Sample Mendelian Randomization (TSMR) is an analytical approach that uses genetic variants as instruments to estimate the causal effect of an exposure on an outcome. This method has the power to mitigate confounding and reverse causation, two common issues in observational studies, by leveraging the random allocation of alleles at conception. Thus, TSMR can provide more reliable evidence of a causal association than traditional epidemiological studies (10).

Despite advances in understanding the genetic basis of these diseases individually, there remains a gap in knowledge regarding the causal relationships between celiac disease and type 2 inflammatory diseases. Previous research has often been limited to observational studies that cannot determine directional causality (11,12). Our study aims to address this gap by employing TSMR to explore the potential genetic associations between celiac disease and type 2 inflammatory diseases, thereby enhancing our comprehension of their shared etiological pathways and contributing to a more integrated approach to their diagnosis and management.

## Methods

### Data Sources

Our study sourced genome-wide association study (GWAS) datasets for celiac disease, along with asthma, allergic rhinitis, and atopic dermatitis, from the MRBase platform (13). These comprehensive case-control studies focused primarily on individuals of European ancestry to ensure a genetically homogeneous sample for effective statistical analysis.

#### Instrumental Variable Selection

Instrumental variables (IVs) were selected from single nucleotide polymorphisms (SNPs) that reached genome-wide significance (P < 5.0×10^-8). To curtail bias due to linkage disequilibrium (LD), SNPs were clumped with an r^2 < 0.001 within a 10,000 kb window. The selected IVs from the GWAS datasets for each health outcome were rigorously documented, detailing effect alleles, betas, standard errors, and p-values. The strength of each IV was quantified by the F-statistic, with values exceeding 10 signifying adequate instrument strength.

#### Research Design Assumptions

Our Mendelian Randomization (MR) analysis was predicated on three critical assumptions:

1. The IVs are significantly associated with the exposure (celiac disease).
2. The IVs are not associated with any confounders of the exposure-outcome relationship.
3. The IVs affect the outcomes exclusively through their impact on the exposure.

## Statistical Analysis

In our Mendelian Randomization (MR) analysis, we harmonized SNP effects on celiac disease and type 2 inflammatory diseases using MRBase, ensuring allele consistency. Our Two-Sample MR approach encompassed multiple methodologies: the primary inverse-variance weighted (IVW) method integrated SNP data to estimate causal effects, while the weighted median, MR-Egger, simple mode, and weighted mode analyses provided supplementary insights, including checks for horizontal pleiotropy. Sensitivity and the robustness of our results were vetted through Leave-One-Out and MR Steiger tests, the latter confirming the temporality of the genetic relationship. Incorporating the MR-PRESSO tool allowed us to detect and adjust for outliers. Power calculations assessed our sample’s sufficiency, outlined by Brion et al. (14) (Available on: https://shiny.cnsgenomics.com/mRnd/). All statistical procedures were performed via MRBase web application (13), and R (Version 2023.03.0+386), utilizing the TwoSampleMR and MR-PRESSO packages, using an alpha of 0.05 to define statistical significance.

## Results

### Genetic Instrumentation and Sample Characteristics

In our Mendelian Randomization (MR) analysis, we used 15 genetic instruments from GWAS data to investigate the associations between celiac disease (ID: ieu-a-1058, Sample size: 24,267) and three type 2 inflammatory diseases: atopic dermatitis, asthma, and allergic rhinitis. The GWAS dataset for atopic dermatitis (ID: ebi-a-GCST90027161, Sample size: 796,661) included 16,121,213 SNPs, reported by Sliz E et al., 2021 (15). Asthma (ID: ebi-a-GCST90018795, Sample size: 449,500) and allergic rhinitis (ID: ebi-a-GCST90038664, Sample size: 484,598) datasets were obtained from the studies by Sakaue S et al., 2021 (16), and Dnerta HM et al., 2021 (17), respectively.

### Association Analyses

#### Celiac Disease and Atopic Dermatitis

Using the Inverse Variance Weighted (IVW) method, we identified a significant positive genetic association between celiac disease and atopic dermatitis. The OR was calculated to be 1.037 (95% CI: 1.015 - 1.059), suggesting a modest increase in the risk of atopic dermatitis with celiac disease. The original β_IVW was 0.0360474 with a standard error of 0.0109833 (p-value = 0.0010306), indicating a statistically significant association (Figure 1 and 2). The MR Egger intercept did not indicate pleiotropy (p-value = 0.5450277), although moderate heterogeneity was observed (Q = 27.58020, p-value = 0.0161667). Power analysis using the two-stage least squares method, with the results from the IVW model, indicated a power of 62%, with an NCP of 5.17 and a high F-statistic of 4765.56. MR-PRESSO analysis supported this association, showing a moderate causal estimate (β = 0.03604742, SE = 0.010983291, p = 0.0054539057). Although the MR-PRESSO Global Test suggested the presence of outliers (p = 0.033), the distortion test indicated that these outliers (indices 8 and 14) did not significantly alter the causal estimate (p = 0.246).

**Figure 1.**
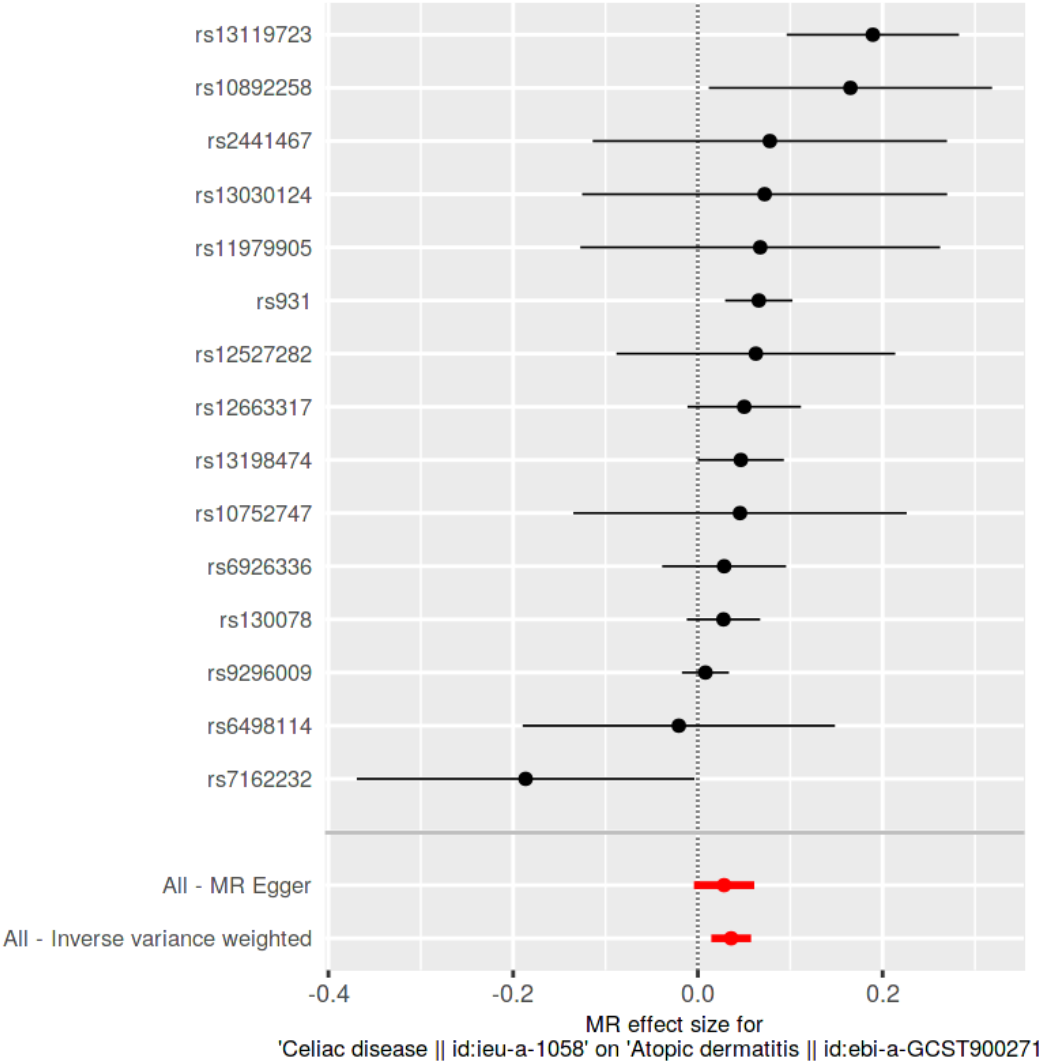
Scatter Plot of SNP Effects on Atopic Dermatitis via MR Egger and Inverse Variance Weighted Analyses.

**Figure 2.**
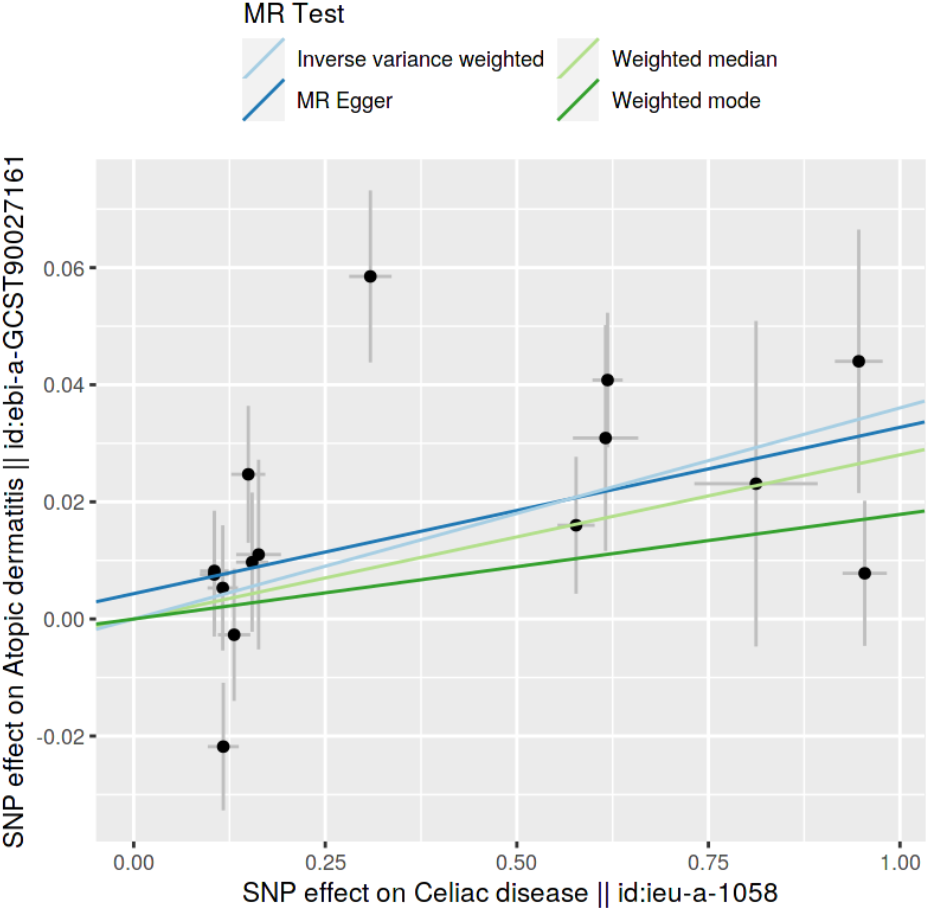
Comparison of SNP Effect Sizes on Atopic Dermatitis Across Different MR Methods.

#### Celiac Disease and Asthma

Our MR analysis suggested a slight protective effect of celiac disease against asthma, with the Weighted Median method yielding an OR of 0.97 (95% CI: 0.96 - 0.98). The original β was - 0.0255837 with an SE of 0.0079101 (p-value = 0.0012193), showing a negative association. However, the MR Egger method found no significant association (β = -0.0066664, p-value = 0.7849596), and high heterogeneity was noted in the results (Q = 120.2595, p-value = 0), with no evidence of pleiotropy from the Egger intercept (p-value = 0.5845075) (Figure 3 and 4). Power analysis based on the Weighted Median method using the two-stage least squares approach yielded a power of 0.36, with an NCP of 2.60 and a robust F-statistic of 4765.56, indicating moderate power to detect the association between celiac disease and asthma. The MR-PRESSO analysis showed a non-significant causal estimate (β = 0.003236846, SE = 0.01574856, p = 0.8401144) with significant heterogeneity (Global Test p < 0.001). The distortion test (p = 0.707) indicated that identified outliers did not significantly affect the result.

**Figure 3.**
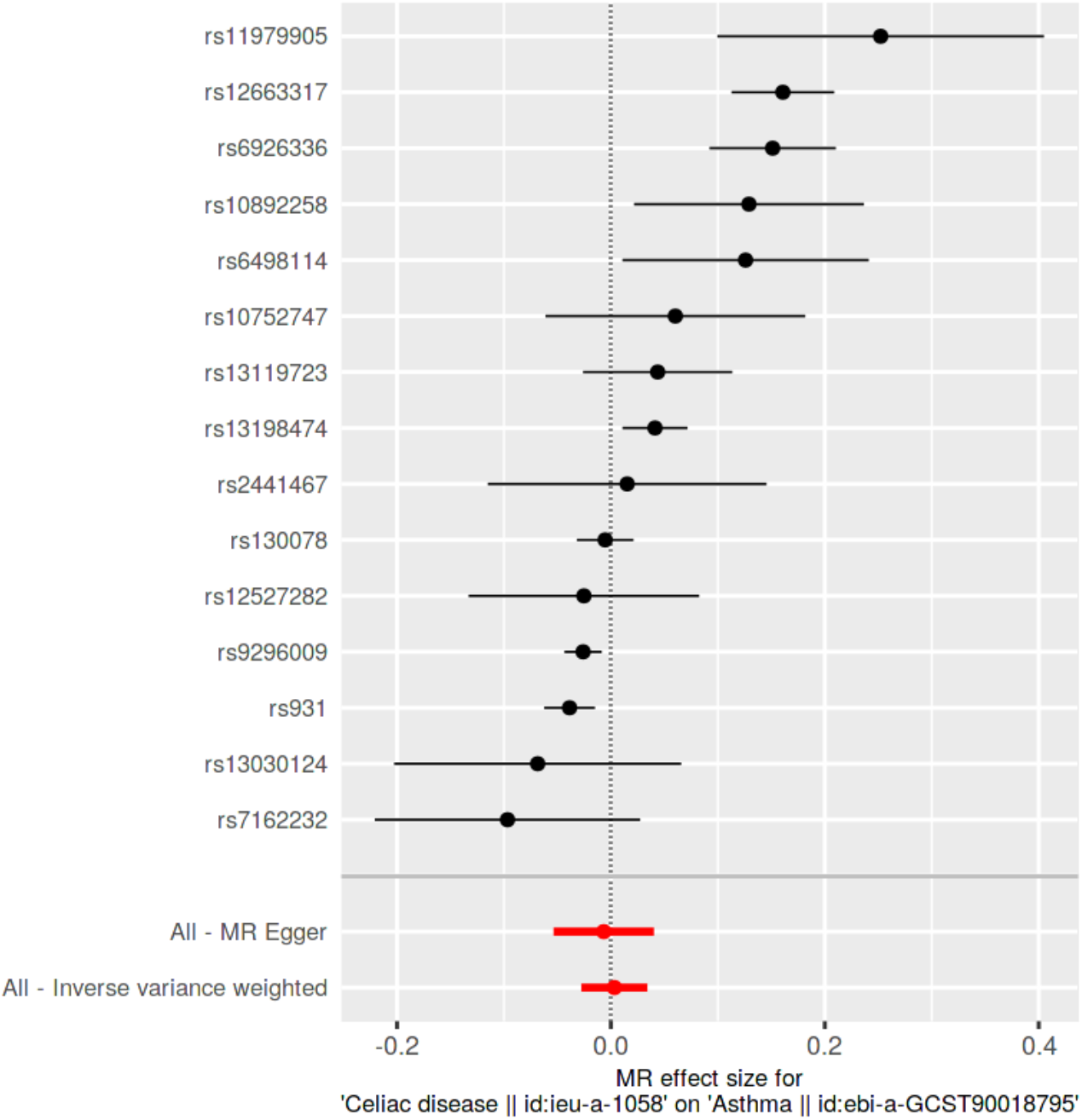
Scatter Plot of SNP Effects on Asthma via MR Egger and Inverse Variance Weighted Analyses.

**Figure 4.**
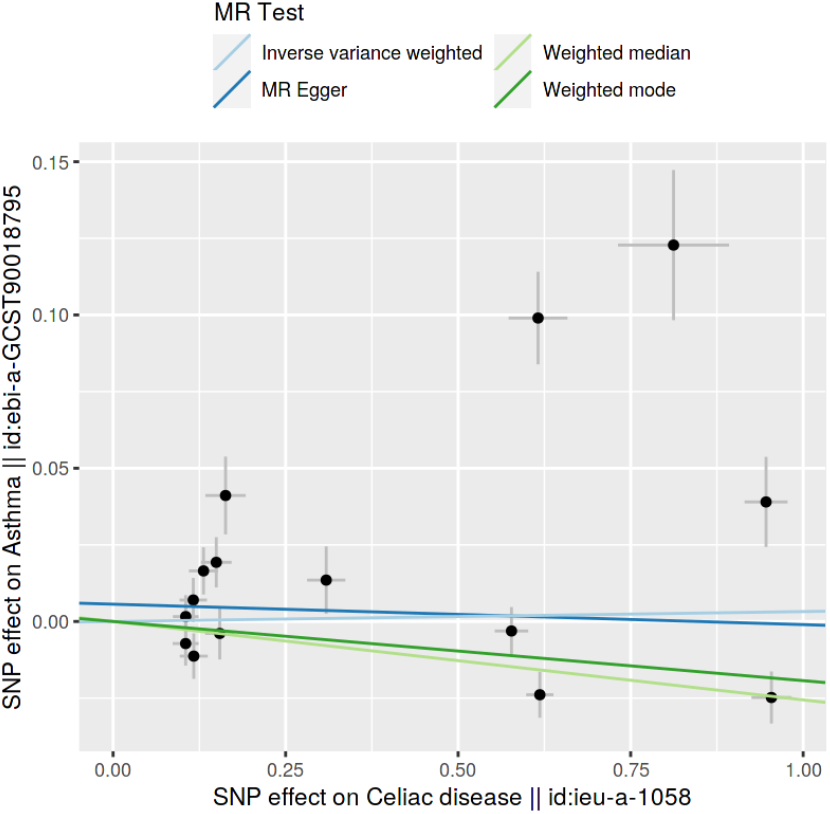
Comparison of SNP Effect Sizes on Asthma Across Different MR Methods.

#### Celiac Disease and Allergic Rhinitis

There was a significant but very small association between celiac disease and allergic rhinitis, as demonstrated by an IVW method-derived OR of 1.002 (95% CI: 1.0004 - 1.0032). The original β_IVW was 0.0018132 with an SE of 0.0007119 (p-value = 0.0108604). Notable heterogeneity was present (Q = 57.33469, p-value = 3e-07), and the MR Egger intercept did not indicate pleiotropy (p- value = 0.6265305) (Figure 5 and 6). Power analysis for the IVW model indicated a moderate power of 0.21, with an NCP of 1.31, despite the strong instrument strength (F-statistic = 4765.56). The results were supported by MR-PRESSO analysis (β = 0.001813204, SE = 0.0007118537, p = 0.02324515). Despite heterogeneity (Global Test p < 0.001), the distortion test (p = 0.715) indicated the minimal influence of outliers (indices 2, 9, 14) on the overall estimate.

**Figure 5.**
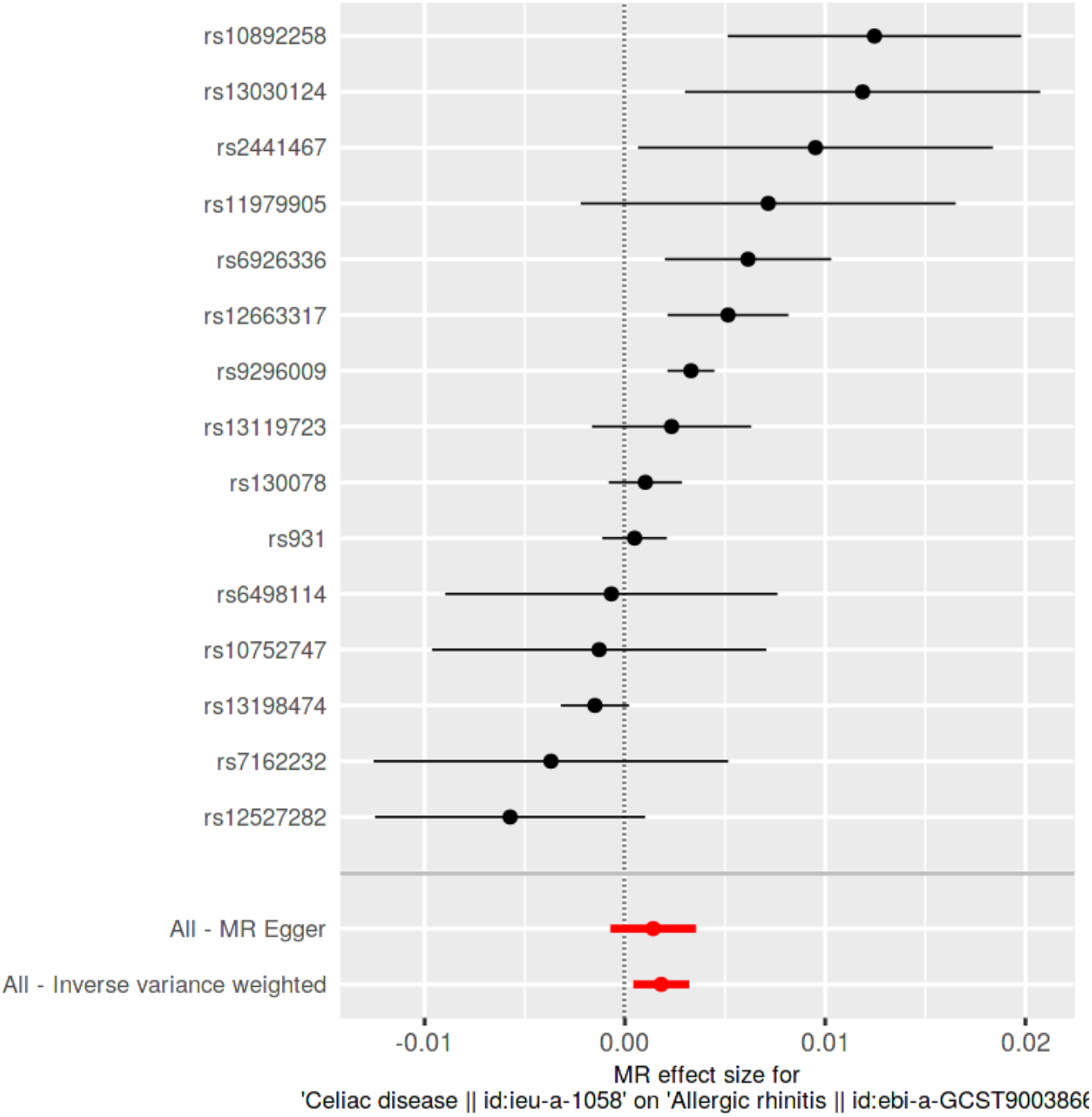
Scatter Plot of SNP Effects on Allergic rhinitis via MR Egger and Inverse Variance Weighted Analyses.

**Figure 6.**
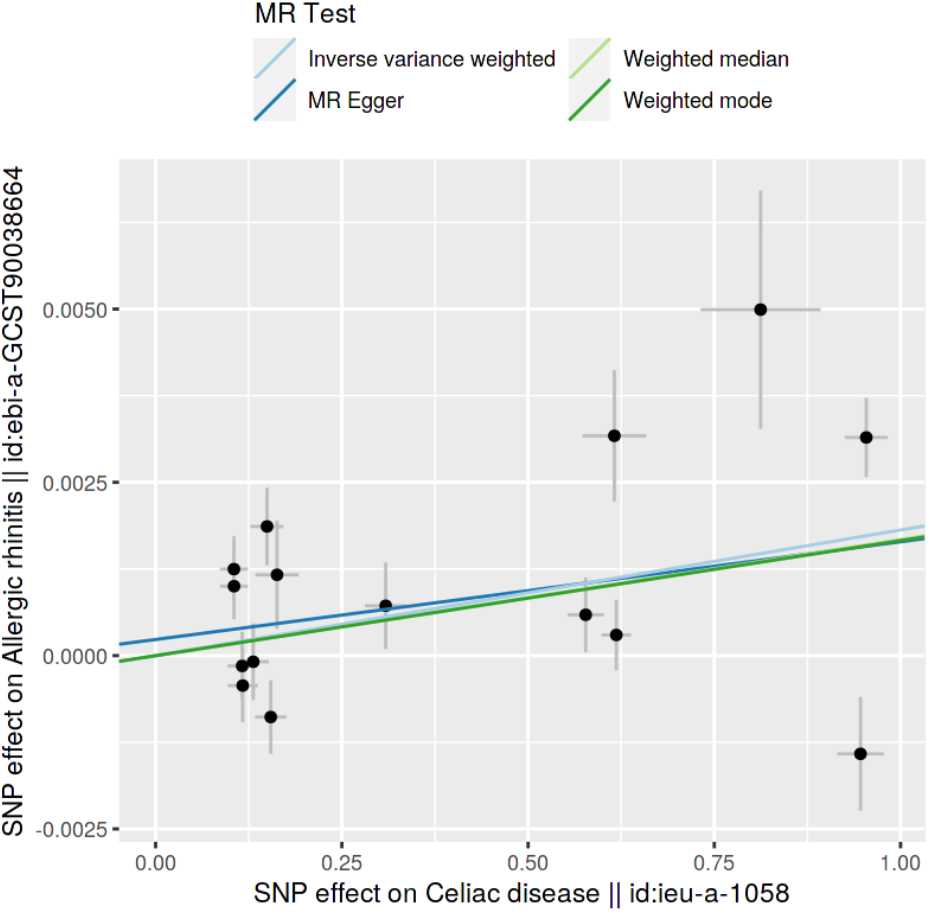
Comparison of SNP Effect Sizes on Allergic rhinitis Across Different MR Methods.

#### Causal Directionality Testing

The causal directionality of the exposure-outcome relationship was assessed using the MR Steiger approach. The test confirmed that the genetic variants associated with celiac disease are indeed upstream of the outcomes, ensuring that the relationship is not due to the outcomes influencing the risk for celiac disease. The Steiger p-value was effectively zero for all three outcomes, reinforcing the direction of causality from celiac disease to each of the type 2 inflammatory diseases.

#### Sensitivity and Publication Bias Assessments

Leave-one-out sensitivity analyses ensured the stability of our findings, as the exclusion of individual SNPs did not significantly alter the results for any of the diseases studied (Figure 7). Furthermore, funnel plots exhibited symmetrical distributions of effect sizes across all outcomes, providing no evidence of publication bias (Figure 8).

**Figure 7.**
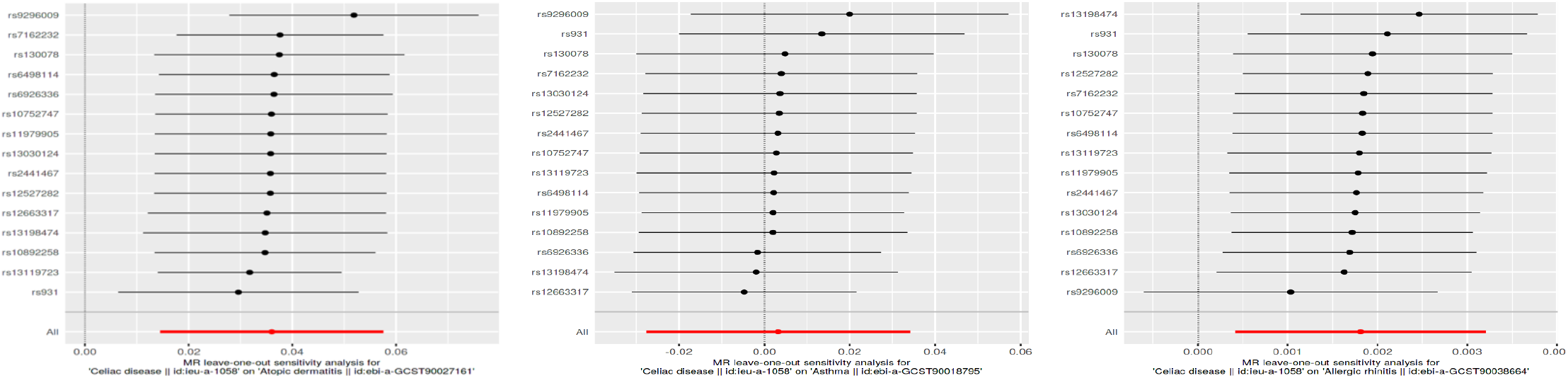
Leave-One-Out Sensitivity Analysis for SNP Effects on Atopic Dermatitis, Asthma, and Allergic Rhinitis.

**Figure 8.**
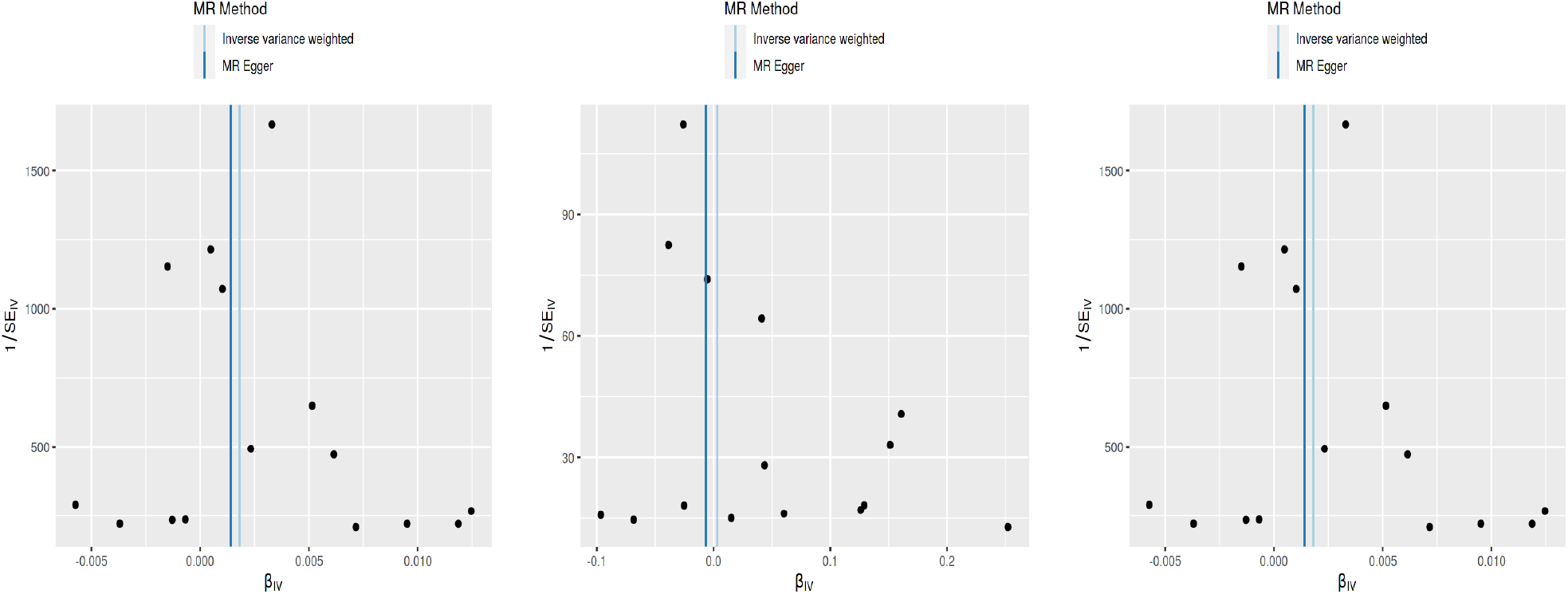
Funnel Plots for Assessment of Publication Bias in MR Analyses.

## Discussion

### Summary of Research and Results

In our research employing Two-Sample Mendelian Randomization (TSMR), we investigated the genetic links between celiac disease and three type 2 inflammatory diseases: atopic dermatitis, asthma, and allergic rhinitis, using 15 genetic instruments. The findings revealed a modestly increased risk of atopic dermatitis (OR = 1.037) and a slight but significant association with allergic rhinitis (OR = 1.002), while indicating a potential protective effect against asthma (OR = 0.97) in the weighted median test. The MR Steiger test validated the causal directionality of these associations, with genetic variants associated with celiac disease preceding the inflammatory diseases. Rigorous sensitivity analyses, including Leave-One-Out tests, upheld the validity of our findings, with no publication bias evident in funnel plot assessments. However, despite a strong instrument strength, the power to detect associations for asthma and allergic rhinitis was limited.

The MR-PRESSO analysis underscored the robustness of the atopic dermatitis and allergic rhinitis associations but advised caution in interpreting the asthma results due to outlier influence and heterogeneity.

### Interpretation of Main Results and Their Implications

In interpreting the main results and their implications, it is crucial to consider the epidemiological link between celiac disease (CD) and type 2 inflammatory diseases. While our study primarily delves into the genetic underpinnings of this association, previous epidemiological studies have laid the groundwork for understanding the clinical coexistence of these conditions. For instance, Karhus et al. identified a potential association between CD and IgE sensitization to various allergens, although subsequent replication studies have yielded inconsistent results (18). Similarly, Majasiak et al.’s systematic review, the first of its kind, underscored the possibility of concurrent allergic IgE-mediated allergy (A-IgE) and CD. This review highlighted cases where CD patients exhibited A-IgE to allergens, particularly wheat, leading to symptoms like atopic dermatitis and anaphylactic shock, even after adopting a gluten-free diet (19). These epidemiological insights, mainly the association of celiac disease with autoimmune disorders and allergies (20,21), reinforce the significance of our genetic analysis, suggesting that the observed genetic links may reflect a broader, clinically relevant interaction between CD and type 2 inflammatory diseases.

The observed associations between celiac disease and type 2 inflammatory diseases in our study provide valuable insights into the shared genetic pathways underlying these conditions. The positive association with atopic dermatitis suggests a common genetic predisposition towards an exacerbated inflammatory response, possibly mediated through shared immunological pathways (22,23). The possible protective effect against asthma highlights a more complex interplay, where certain genetic factors associated with celiac disease may confer a reduced risk for asthma (24), potentially through differential immune regulation (25). The significant association with allergic rhinitis further underscores the genetic linkage between celiac disease and allergic conditions. These findings suggest that the genetic predisposition to celiac disease may extend beyond the traditionally understood gastrointestinal manifestations, influencing broader systemic inflammatory processes.

Our study, utilizing TSMR, offers new insights into the genetic causality between celiac disease and type 2 inflammatory diseases, moving beyond the observational correlations highlighted in prior research (5,11,12,18–21,24). Specifically, our findings reinforce the epidemiological evidence for shared genetic pathways in the inflammatory processes of celiac disease and type 2 inflammatory diseases (2,9,12). For instance, the genetic predisposition to an exaggerated inflammatory response in celiac disease and its positive association with atopic dermatitis may indicate overlapping pathways in Th2-mediated immunity (11). On the other hand, the possible protective association with asthma suggests a complex interplay where certain genetic components in celiac disease might modulate inflammatory responses differently, possibly through immune regulatory mechanisms (26). Understanding these shared pathways can lead to more comprehensive models of disease pathogenesis.

### In-Depth Discussion of the Literature Insights and Study Findings

Our study’s insights into the genetic associations between celiac disease and type 2 inflammatory diseases intersect with existing literature, particularly regarding the role of pro-inflammatory cytokines and immunomodulatory activities identified in celiac disease due to gliadin sequences (27,28). The positive association with atopic dermatitis observed in our findings may reflect these shared inflammatory pathways, potentially involving Th2-mediated immune responses (22). Furthermore, the significance of HLA region variants in celiac disease, as demonstrated in previous GWAS studies (23,29,30), parallels our observations of genetic overlap, especially considering the crucial role of HLA alleles in immune response and their associations with atopic dermatitis and asthma (23,31).

Additionally, the association of specific HLA Class I allelic variations with atopic dermatitis (29), aligns with our findings, suggesting a complex genetic interplay in these diseases. The slight, but nevertheless possible protective effect against asthma in our study could hint at differential genetic interactions within the HLA super-locus, a region known for its involvement in various asthma phenotypes (31). This aligns with the heterogeneity seen in asthma’s genetic studies and the difficulty in studying class II genes due to human population diversity and the complexity of the MHC (31,32). Furthermore, the involvement of regulatory T cells (Tregs) in both celiac disease and asthma (25,33), might explain the complex relationship we observed, suggesting potential commonality in the regulatory pathways of immune response. This said, the link between asthma and celiac disease necessitates deeper exploration in forthcoming studies, particularly due to the inconclusiveness of numerous tests (such as MR Egger and IVW). While a minor protective influence is conceivable, it cannot be conclusively affirmed nor dismissed, as the weighted median method can offer a more accurate estimate in some cases (34). Concerning allergic rhinitis, the connection between HLA-DQB1 gene polymorphisms and allergic diseases reported by Cardaba et al. and Tokunaga et al. (35,36) reinforces our findings of a possible significant relationship between celiac disease and allergic rhinitis, highlighting intricate genetic landscapes shared among these conditions.

### Strengths and Limitations of the Study

One of the principal strengths of our study lies in the application of Two-Sample Mendelian Randomization (TSMR), which offers a robust approach to infer causal relationships by addressing common issues of confounding and reverse causation often encountered in observational studies (10). The use of extensive GWAS datasets and a variety of MR methods (IVW, MR Egger, weighted median) to ensure reliable results, further validated by sensitivity analyses and the MR Steiger test (37). However, limitations include the primary use of European ancestry data, potentially limiting wider applicability, and the possibility of residual pleiotropy (38). Additionally, while we focused on genetic associations, this doesn’t encompass all factors, such as environmental or lifestyle influences, impacting these diseases. Therefore, our findings contribute to, but do not fully elucidate, the complex genetics of type 2 inflammatory diseases.

### Conclusion and Future Research

In conclusion, our study provides significant insights into the genetic associations between celiac disease and type 2 inflammatory diseases, highlighting a potential shared genetic foundation. While our findings are a crucial step towards unraveling the complex interplay between these conditions, they represent just one piece of a larger puzzle. Our research underscores the need for more in-depth investigations into the shared pathways of these diseases, which could potentially lead to improved understanding and treatment strategies, with more targeted clinical interventions and personalized treatment approaches for patients with celiac disease and related inflammatory disorders. Future research should focus on including more diverse populations to enhance the generalizability of the results. Additionally, exploring the environmental, lifestyle, and epigenetic factors that interact with these genetic associations will be essential for developing a more comprehensive understanding of these diseases.

## Data Availability

All data produced in the present study are available upon reasonable request to the authors

## Acknowledgment

none

